# Implementation of dementia communication skills training in acute hospitals: a longitudinal, mixed-methods case study evaluation

**DOI:** 10.1101/2025.07.02.25330714

**Authors:** Rebecca O’Brien, Danai Theodosopoulou, Marie Janes, Rachel Clark, Rowan H. Harwood, Andrew Papworth, Suzanne Beeke, Louise Bramley, Aquiline Chivinge, Sarah Goldberg, Sue Haines, Alison Pilnick, Kate Sartain, Morag Whitworth, Claire Surr

**Author notes:** Corresponding author address and e-mail, Claire Surr, Centre for Dementia Research, School of Health, Leeds Beckett University, Leeds, LS1 3HE, UK. Note: O’Brien and Surr are joint lead authors.

## Abstract

**Purpose:** People with dementia occupy c.25% of acute hospital beds. Being in hospital can lead to distress in people with dementia, exacerbated by the difficult acute hospital environment and lack of staff preparedness to meet their often-complex needs. Acute hospital staff identify supporting distressed patients with dementia as a practice challenge. Communication skills training can improve interaction quality and staff confidence, however little communication research to date has been conducted in acute hospitals. This study aimed to implement a communication skills training programme for preventing and responding to distress in patients with dementia in acute hospitals, and to assess its impact.

**Methods:** A longitudinal, mixed-methods, multiple case study design was employed in six wards across three English acute hospitals. The Kirkpatrick training evaluation framework (reaction to training, impact on knowledge and confidence, behaviours and outcomes for patients and staff) underpinned the study, with qualitative observational and interview, and quantitative survey and outcomes data collected immediately pre, post and 1-3 months post training. The [redacted] communication skills training programme included three communication trainables identified in an earlier conversation analysis study phase. It was delivered over two half-days, by local clinical educators, upskilled via a train-the-trainer programme.

**Findings:** A total of 145 staff attended at least one half-day of training. Delivery was feasible, however practical challenges with training organisation and freeing up staff to attend occurred. Staff who found it engaging and relevant to their role, valued its interactive content and the opportunity for reflection and implementation of skills between sessions. A statistically significant increase was found pre- to immediate post-training on staff communication knowledge (CI 0.01-1.2) and confidence in caring for people with dementia (CI 4.9-7.3). Staff reported a range of areas of learning aligned to the trainables. Over 90% of staff said they planned to implement the training in practice and many provided concrete examples of application. Challenges in applying the specific communication principals taught to wider practice situations were identified by some staff. Impact on observed patient agitation levels and staff communication practices were challenging to evidence objectively in the context of the acute hospital environment.

**Conclusion/original contribution:** It is feasible, although challenging, to deliver empirically based communication skills training to support acute hospital staff to better care for patients with dementia who may become distressed. It can lead to perceived increases in knowledge and confidence to support distress in this population.

**Study registration:** Not registered

## Background

At least 1 in 4 UK acute hospital beds are occupied by people living with dementia(1) usually receiving care for a condition additional to their dementia; a figure which is reflected across other high-income countries(2, 3). Communication difficulties are a common symptom of dementia including expression (e.g. word finding difficulties, reduced verbal fluency) and problems understanding others (e.g. the content and meaning of what is said, following instructions)(4). Given this, acute hospitals are difficult environments for people with dementia, with many structural issues(3) making it challenging for their often complex needs to be appropriately understood and met(5). These include inadequate staff training on dementia, busy and disorientating physical environments, and inflexible care processes and routines that do not consider the unique needs of people with dementia(6, 7). In response people with dementia may exhibit behaviours such as agitation, distress and repetitive calling out, may engage in exit seeking or resist personal and medical care, as a way of expressing their inadequately met needs(8). These behaviours can be further escalated by the unsupportive physical and social environment and poor quality of staff interactions (7) leading inappropriate responses to their distress, and resulting in an escalating cycle that can lead to poor outcomes for the person with dementia. This includes longer length of stay, increased rate of functional decline and increased likelihood of requiring long-term care(7, 9). It is therefore unsurprising that many acute hospital staff report caring for people with dementia, particularly those who are distressed or likely to become so, to represent a significant challenge they often feel unprepared to meet(10).

Psycho-social and environmental interventions are the recommended first line approach to management of agitation and distress in people with dementia (11), with person-centred care central to this (12). While acute hospital staff generally recognise the need to deliver person-centred care(10), understanding specifics of *how* to do this successfully in daily practice can remain elusive(13). Improved dementia training is consistently identified as underpinning staff ability to deliver person-centred care (10). Communication skills training in dementia care has been found to improve interaction quality and increase staff knowledge, skills and competency(14–16). However, evidence of impact on outcomes for people with dementia remains limited(16) and more research is needed on what communication approaches work, in which situations(17). To date, little of the dementia communication research has been conducted in acute hospital settings, and few dementia training interventions in acute hospital settings have focussed on communication skills(16, 18–20). Thus, knowledge about effective communication approaches in this unique context remains limited.

In an earlier phase of this study, we video/audio recorded naturally occurring healthcare interactions (n=53) between healthcare practitioners and patients with dementia who were prone to or in distress, in two English National Health Service hospitals. We used Conversation Analysis to identify communication approaches that worked to avoid, de-escalate or resolve any distress, which are reported in separate papers [citations redacted]. Conversation Analysis (21) is a research method with the ability to uncover, and make explicit, tacit communication practices. It uses detailed and rigorous study of video or audio recordings of naturally occurring interactions to enable the identification of verbal and non-verbal communication practices which would otherwise have remained unconscious (in the sense of inarticulable) to the participants. Conversation Analysis analyses what people actually do when communicating, rather than what they think or say they do, and can be used to reveal what is interactionally successful in particular contexts(21). It has been used to identify features of successful communication in healthcare(22) and to develop authentic communication skills training to improve delivery of health and social care, including for people with dementia(23–25). In summary, this analysis identified approaches acute hospital staff use to deal successfully with interactional challenges such as not being able to meet a patient’s expressed need, the patient being in a different reality, and the unavoidable need to undertake health and care tasks with patients that could cause them or are causing them distress (such as giving them an injection or repositioning them in bed).

These approaches formed the basis of three ‘trainables’ (following interactional rules [citation redacted]; managing different realities [citation redacted]; accounting for care [citation redacted]) with two or three communication practices attached to each. These formed the basis of the [redacted] communication skills training. How these trainables were developed into a training programme is discussed below.

### Aims

This study aimed to implement the [redacted] dementia communication skills training programme for acute hospital staff and assess its impact.

It addressed the following research questions:

1. Can the [redacted] communication skills training programme be implemented in practice in a range of acute hospitals, and what are the barriers to and enablers of this?
2. What is the impact of the [redacted] dementia communication skills training programme on acute hospital staff knowledge, confidence, practices and on outcomes for patients and staff?

## Methods

### Design and theoretical position

A longitudinal, mixed methods, multiple, embedded case study design was employed following Yin(26).

Kirkpatrick’s (27, 28) four-level model for the evaluation of training interventions underpinned study design, methods and analysis including evaluation of reaction to the training, its impact on learning (i.e. knowledge, attitudes and confidence), staff behaviours and outcomes for patients and staff. The COM-B model (29) for understanding behaviour change was utilised to explore training implementation.

### Setting

Three English NHS acute hospitals (Sites A, B and C) took part in the study, one in the south, one in the midlands and one in the north of England. Each hospital represented a case. Two wards delivering care to older people were identified by each participating site to take part as embedded cases.

### [REDACTED] training programme

The [REDACTED] training intervention was developed through four half-day, in-person workshops with a group of up to 20 expert stakeholders, including clinicians (physical and mental health nurses, doctors, speech and language therapists, healthcare assistants), experts in healthcare education and digital learning and patient and public involvement representatives. Workshops included presentation and discussion of the current evidence base on impactful dementia training (30–32), the three trainables of communication approaches that form the core of the programme, identified in the phase one Conversation Analysis study [citations redacted], evidence-based communication skills resources (33, 34) and digital learning.

In workshop four, the TIDieR framework(35) for intervention description, informed by summaries from the previous workshops, was used to guide decisions including: who would deliver the training; duration, pattern and mode of delivery; and key content, resources and training techniques.

The resulting training programme was interdisciplinary and interactive, including multi-media PowerPoint presentations with teaching notes, video resources including clips from the phase one Conversation Analysis study that we had participant consent to use for training purposes, and skills practice. It was delivered face-to-face by dementia-experienced clinical educators, over two half days, separated by a reflective period of 2-4 weeks.

A pilot programme was delivered by researchers to nine staff including doctors, nurses, an Occupational Therapist and a Speech and Language Therapist. Four were from one of the participating sites but in roles that would not involve delivering care on the participating wards, and five were from neighbouring NHS organisations. Feedback from pilot participants helped to refine the final training materials prior to implementation.

### Participants

#### Clinical Educators

Clinical educators were three experienced staff from each hospital site, who had a remit for dementia education within their role, who were able and willing to attend a train-the-trainer programme, and then onward deliver the communication training to practitioners in their hospital. The train-the-trainer programme consisted of two full days training separated by one month. The morning sessions involved receiving the [REDACTED] half-day content and the afternoons included training on delivery methods, with time for practice, problem solving and planning.

#### Health Care Practitioners

We aimed to train c.50 Healthcare professionals per case study site on the [REDACTED] programme, including 50% of staff working on each ward, to achieve a ‘critical mass’ of trained staff.

Healthcare professionals recruited to the study were all staff who worked in one of the hospital sites and who delivered care to patients on one of the participating wards. This could include staff in any direct care delivery role and staff (for example Allied Health Professionals and doctors) who may work across multiple wards. Inclusion criteria were that staff were willing to attend the training, complete the pre- and post-training data collection and potentially be approached to undergo observations and interviews.

#### Ward managers

The ward manager or deputy from each participating ward.

#### Patients, family members and informal carers

Patients accommodated on one of the participating wards during the study period, and their associated family/carer(s). They needed a recorded diagnosis of dementia and to be prone to distress as confirmed by the clinical team. Patients were excluded if they were deemed by the clinical team as being too unwell to participate, or if they lacked mental capacity to give informed consent and researchers were unable to identify a personal consultee who could provide advice on their wishes.

### Recruitment

#### Staff participants

Clinical educators were identified and approached by their organisation’s lead investigator. Ward Managers were approached by the researcher, following their agreement for their ward to participate in the study. Healthcare professionals were initially approached by the clinical educators or the ward manager. Written informed consent was then obtained via a researcher using paper or digital consent forms.

#### Patients, families and carers

The initial approach to patients, and their family members/carers, was via a member of the patient’s regular care team, who gained consent to introduce the researcher. Study explanation and informed consent was undertaken by the researcher. Where patients with dementia lacked the capacity to give informed consent, advice on their wishes around participation was gained via a personal consultee (family member or friend) in line with the English Mental Capacity Act (36).

### Data collection

Data were collected at various time points, from different participant groups. Collected data is summarised in Table 1, alongside the Kirkpatrick evaluation level it provided data for.

**Table 1:** Summary of data collected, and the Kirkpatrick level addressed.

| **Data collected and Kirkpatrick (K1) level addressed** | **Pre training (T0)** | **During training** | **Immediately post training (T1)** | **Within 2 weeks of day 2 training** | **1-3 months post training (T2)** |
| --- | --- | --- | --- | --- | --- |
| **Questionnaire containing the following components** |  |  |  |  |  |
| **Demographics** | CE, HCP |  |  |  |  |
| **Programme evaluation questionnaire (K1)** |  |  | CE, HCP |  | HCP |
| **Communication knowledge in dementia test (K2)** | CE, HCP |  | CE, HCP |  | HCP |
| **Confidence in dementia scale (K2)** | CE, HCP |  | CE, HCP |  | HCP |
| **Confidence in delivery of training questionnaire (K1)** | CE |  | CE |  | CE |
| **Other data** |  |  |  |  |  |
| **Semi structured interviews (K1, 2, 3, 4)** |  |  |  | CE | CE, HCP, WM, P, F/C |
| **Training delivery data (number of sessions, staff attending etc)** |  | CE |  |  |  |
| **Observations of training delivery (K1, 2, 3)** |  | R |  |  |  |
| **Practice observations of agitation (K4)** | R |  |  |  | R |
| **Practice observations of communication behaviours (K3)** | R |  |  |  | R |

CE = clinical educator, HCP = health care professional, WM = ward manager, P = patient, F/C = family member/carer, R = collected by researcher

#### Programme evaluation questionnaire

A bespoke questionnaire with Likert scale (1-10 with a higher score indicating greater agreement/satisfaction) and open-ended questions covering training design, delivery and satisfaction.

#### Communication knowledge in dementia test

A 34-item, multiple-choice measure developed by the research team based on the learning outcomes of the programme. We developed similar questionnaires for a previous communication programme [5,49]. A higher score indicated greater communication knowledge.

#### Confidence in Dementia Scale (37)

A 9-item measure assessing self-efficacy to care for people with dementia, measured on a 5-point Likert scale, with a higher score indicating greater confidence.

#### Confidence in delivery of training questionnaire

An 11-item questionnaire measured on a 10-point Likert scale, with a higher score indicating greater confidence, developed by the research team in a previous study for evaluating a train-the-trainer programme [redacted]. The questionnaire includes items designed to evaluate confidence levels in various aspects of delivering the [redacted] training programme. Specifically, confidence in facilitating [redacted] workshops, explaining the evidence base behind [redacted], addressing questions from healthcare professionals regarding the programme, teaching the three sets of ‘trainables’, organising the programme at the hospital, ensuring adequate time for programme delivery, and the belief that Healthcare professionals will find the programme valuable and useful.

#### Semi-structured interviews

These were conducted by a researcher either in-person or online using video conferencing software, and were audio recorded and transcribed verbatim. Interview topic guides were developed in collaboration with staff and lived experience stakeholders.

#### Training attendance

Clinical educators recorded how many training sessions they delivered, and which staff had attended training.

#### Training observations

Researcher observations of two half-day [REDACTED] sessions (constituting one full programme overall), at each site, using a framework based on one developed for previous research(32), covering: Training content and delivery; learners’ reactions, evidence of learning, intentions in relation to changes in their future practice and the way they thought this might impact on quality of care; and observed barriers and facilitators to learning.

#### Practice observations – communication behaviours

A researcher conducted non-participant written observations of one or more consenting healthcare professionals who were due to (pre training)/had attended (post-training) the [REDACTED] training, using a checklist developed by the research team, based on the [REDACTED] trainables, and a checklist previously developed to assess communication skills training (18, 38). The checklist contained seven positive and three negative communication practices. HCP interactions with patients with dementia who were prone to become distressed were undertaken for multiple periods of up to 20-minutes per site. Patients were fully anonymised at the point of data collection. A combined total of up to two-hours of observation was conducted per ward at each time point. Data are presented as the mean number of positive and negative communication behaviours observed per 100 minutes of observation.

#### Practice observations - agitation

Researcher non-participant observation of one or more anonymised patients with dementia who were prone to distress, for a period of two consecutive hours at each time point. The frequency of agitated behaviours was assessed using the Cohen-Mansfield Agitation Inventory observational version (CMAI-O) (39). This is a 29-item measure with each item scored on a four-point scale, summed to generate a total agitation score per individual. Higher scores indicate greater frequency of agitated behaviours. The intensity of agitated behaviours was assessed using the Pittsburgh Agitation Scale(40), which includes four domains; each scored on a 5-point Likert scale. A total score per participant is derived with a higher score indicating greater intensity of agitated behaviours. Data are presented as the mean agitation score for the two-hour observation period.

### Data analysis

Data analysis involved the following steps(26)

1. Analysis of each data source/type
2. Integration of the data sources by case (ward, NHS Trust) to produce individual cases of training implementation, drawing on multiple data sources
3. Comparison and integration of data within and cross-case, using data summary tabulation.

Qualitative data were analysed using template variant of thematic analysis (41). Initial deductive codes were developed based on Kirkpatrick’s model, the TIDieR checklist (35) for the reporting of interventions, and the COM-B model (creating a v1 coding template). Familiarisation with nine interview transcripts (3 per case study site), covering a range of participants, was undertaken by six co-authors/researchers, [redacted] and discussed to generate additional inductive codes, in the context of the v1 template. A v2 coding template was subsequently developed and applied by two co-authors [redacted] to all the qualitative data using NVIVO v12(42). Refinements to the template were discussed between the coders as required as coding progressed.

Quantitative analysis was conducted using STATA v18. To assess changes in communication knowledge and confidence, paired-sample t-tests were used to calculate mean scores and assess statistically significant changes pre- to post-training. Frequency distributions were calculated to examine the spread of responses on other programme satisfaction questionnaire items. Changes in observed agitation in patients using the PAS and CMAI-O were compared using independent samples *t*-tests comparing the mean agitation score at each time point. Due to poor return rates at 1-3 months post-training (T2) only baseline (T0) and immediate follow-up (T1) data has been presented.

## Ethical considerations

Ethical approval for this implementation and evaluation study was gained from the National Research Ethics Service [redacted]. All participants gave informed consent ahead of participation. Participants were informed that participation was voluntary and that their decision would not impact their employment, treatment and care, or legal rights. Participants had the option to withdraw at any point. In the case of their withdrawal, the data collected up to that point was retained and used in the final analyses.

## Findings

The findings are presented according to the four Kirkpatrick levels, evaluating training delivery, and barriers and facilitators to this, alongside whether there was evidence of subsequent implementation of skills into practice and associated impact. Table 2 provides a summary of the case study sites.

**Table 2:**
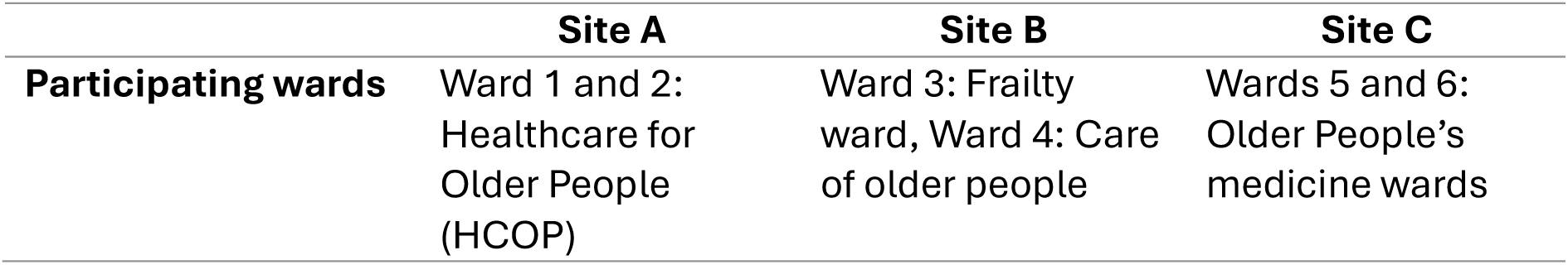
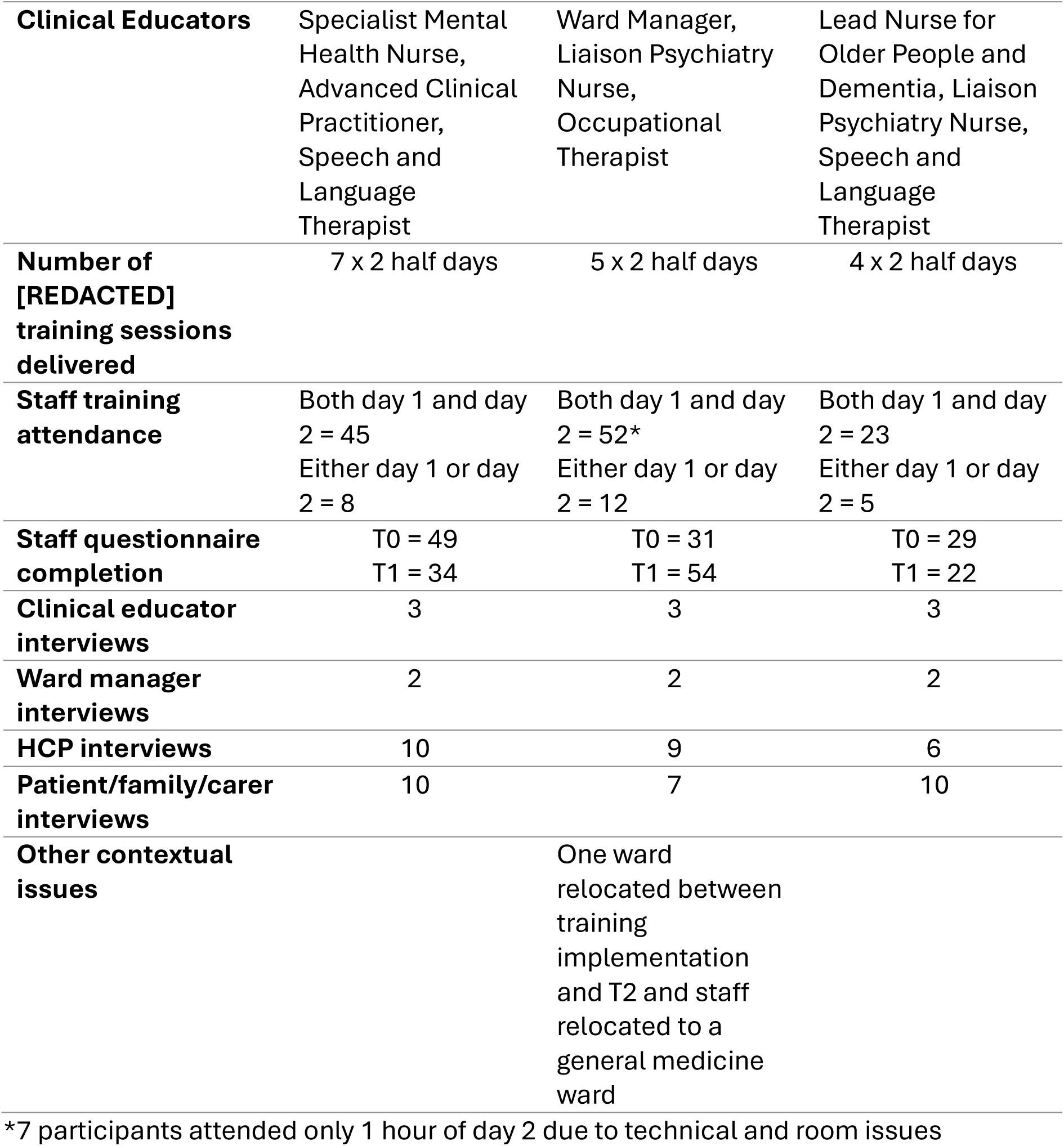
Summary of case study sites.

### Training delivery

A total of 145 staff attended at least one half-day of training (Table 3) across the three sites and 120 completed both days. Training delivery was challenging across all sites given NHS service pressures and the impact this had on organisation’s abilities to release staff for two half days. Sites organised delivery in different ways dependent on the site context e.g. training room availability, how rotas were arranged and how training attendance was backfilled. Only site B achieved the total of 50 staff attending training and in site C only around half of anticipated staff trainee numbers were achieved. Assessing whether 50% of ward staff were trained was complex given staff turnover and ward moves, and the input of a range of staff to care who were not ward-based. In two sites, training for each half day was grouped over a one-week period, whereas in the third site sessions were delivered across a wider timespan.

**Table 3:** Training attendee demographics*.

|  | Site A | Site B | Site C | Total |
| --- | --- | --- | --- | --- |
| **Role n (%)** | 49 (100) | 32 (100) | 29 (100) | 110 (100) |
| Doctor | 2 (4.1) | 0 (0) | 1 (3.4) | 3 (2.7) |
| Nurse | 17 (34.7) | 19 (59.4) | 8 (27.6) | 44 (40) |
| Healthcare Assistant | 10 (20.4) | 8 (25) | 6 (20.7) | 24 (21.8) |
| Allied Health Professional | 14 (28.6) | 3 (9.4) | 7 (24.1) | 24 (21.8) |
| Therapy Assistant | 6 (12.2) | 2 (6.2) | 1 (3.4) | 9 (8.1) |
| Other | 0 (0) | 0 (0) | 6 (20.7) | 6 (5.4) |
| **Gender** |  |  |  |  |
| Female | 37 (75.5) | 20 (62.5) | 25 (86.2) | 82 (74.6) |
| Male | 12 (24.5) | 11 (34.4) | 4 (13.8) | 27 (24.6) |
| Prefer not to say | 0 | 1 (3.1) | 0 | 1 (0.0) |
| **Ethnicity** |  |  |  |  |
| Asian | 20 (40.8) | 8 (25.0) | 10 (34.5) | 38 (34.6) |
| Black | 6 (12.2) | 4 (12.5) | 6 (20.7) | 16 (14.6) |
| Chinese | 0 | 0 | 2 (6.9) | 2 (1.8) |
| Indian | 0 | 0 | 1 (3.5) | 1 (0.9) |
| Mixed | 1 (2.0) | 0 | 0 | 1 (0.9) |
| Sikh | 0 | 0 | 1 (3.5)) | 1 (0.9) |
| White | 20 (40.8) | 20 (62.5) | 6 (20.7) | 46 (41.8) |
| Other | 2 (4.1) | 0 | 3 (10.3) | 5 (4.6) |

\*From T0 survey responses

Clinical educators highlighted some of the challenges and facilitators in training delivery, which included finding time to deliver training irrespective of whether the half day one and two were arranged together as block or distributed over a number of weeks, and the difficulties scheduling two half-days caused for rotas and ensuring all staff were able to attend both sessions. Staff shortages and the need to meet operational demands meant wards were only able to release a small number of staff for each session meaning multiple sessions were needed to attain the proposed critical mass. For most sites this was not feasible to achieve over the delivery period. Having ward managers engaged and supportive of the training ensured the levels of attendance that were achieved.

> *“It was quite hard to organise because it was two separate dates and you had to have the … two weeks in between and then from a nursing perspective, you have to look at all the rosters … and … we had to get the rooms first.”* Clinical Educator Site C
>
> *“We planned the sessions all within one week. All very close together. I think that was an issue because some people, if they were on leave, couldn’t make any of the sessions, whereas if we’d spaced them out a little bit more, we would have probably got better attendance.”* Clinical Educator Site A

Clinical educators noted it was difficult to get staff who might benefit most from the training to attend.

> *“I think some of the ones that came, if you could choose people, they wouldn’t necessarily be the ones you’d have because they’ve already got quite a good knowledge. But it was quite difficult to get some of the others to come.”* Site C Clinical educator

### Reaction to training

There were no substantive differences between reactions to the training across the three sites. The training was evaluated highly positively, with all items on the programme evaluation questionnaire receiving scores of 8+/10 by at least 80% of participants. The only item where there was greater variability of scores was whether participants found the programme challenging where 12% rated it as not challenging (score 1-4), 13% felt neutral (score 5-6), and 76% said it was challenging (score 7-10).

Participants reported the training to be engaging, well-structured and relevant to their role.

> *“It was things that were definitely going to be helping us in the ward, so I think it was just really meaningful. It had a great impact on me as a healthcare assistant.”* Staff participant site A
>
> *“It’s very relatable, because it’s not only in our ward or in our hospital. With the video that they showed, it shows that it also happens in other hospitals, and then it makes us feel like we’re not the only people experiencing this.”* Staff participant site B

The multi-disciplinary nature of the training and interactive elements, such as role play, group discussions, video content using clips from phase one of the study, and scenario-based learning, were particularly valued.

> *“I think it was nice hearing shared knowledge from between the different members of the team. It was quite good that … the training wasn’t just done with occupational therapists all in one room or all nurses … Yeah that … shared learning was really valuable.”* Staff participant site C
>
> *“The opportunities for the discussion … I really liked the video clips and the transcripts … you’d really, like, immerse yourself in the scenario sort of thing ..*.
>
> *This was, like, that is a patient that I’ve seen or … could be like a typical patient. And it was nice to see … the difference that using some of the strategies or not using the strategies made.”* Staff participant site A

The structure of allowing time between the two sessions for reflection and practice application was valued.

> *“I think it’s really helpful that it was divided into two, because first it’s too much if it would have been in one go. It was divided, so we were able to digest, and we were able to apply in our area as well each session.”* Staff participant site B
>
> *“Yeah, I did quite like it because I think they gave us a bit of a task to do and go and reflect. So, I think it was useful to have that little bit of almost like homework a little bit, to like kind of consolidate our learning”* (Staff participant site C)

Less positive aspects of the programme were also identified. The Clinical Educators noted that some staff expected a more prescriptive approach and found identifying ways to apply the examples of communication approaches to a wider range of scenarios in their own practice challenging. This was also evident in the staff interviews.

> *“Some people wanted a bit more of a prescriptive …. So if this happens, we do this, do we? And we were trying to say, well, no, no, we’re just giving you examples so that you can understand what ..the research shown helps. But people I think maybe the training they’ve had previously or the way they’ve been taught is very much [more prescriptive]”.* (Clinical Educator site A)
>
> *“I find we get a lot more of wanting to know, why did I come here, what happened, how long have I been here? Rather than, I want to go home.* {Example in video clip]*”* (Staff participant site C)

When asked about ways to improve the programme reducing perceived repetition of information and accessibility of some of the language used were identified. Participants commented on the use of transcripts of the video communication approaches feeling lengthy, repetitive and complex to engage with.

> *“The transcripts were quite long and repetitive at times.* (Staff survey site A)
>
> *“If the transcripts were just not like an academic transcript would probably help a lot of people.”* (Clinical Educator site C)

Clinical Educators noted some of the language associated with the trainables was not accessible to all staff, without more detailed explanation.

> *“I think there was a bit of language that I think they didn’t quite know .. what we meant. So it took a while to get … that person centred”* (Clinical Educator site C)

Potential improvements included providing programme materials online to improve accessibility and simplified PowerPoint slides.

### Knowledge, attitude and confidence

Scores on the knowledge test were high across all sites at baseline indicating a measurement ceiling effect (Table 4). A small but statically significant increase in knowledge scores between T0 and T1 was found at site A and across combined sites. There was a large and statistically significant increase in confidence in caring for people with dementia at T1 across all sites, also reflected at individual site level (Table 3). Post-training, 88% of respondents felt confident using the communication techniques (scoring 8+/10) and reported their confidence had improved post-training.

**Table 4:** Staff outcome measures data.

| Scale<br>(n= pre, post) | Pre-<br>training<br>(T0) | Post-training<br>(T1) | Mean<br>difference | 95% CI | p-<br>value* |
| --- | --- | --- | --- | --- | --- |
| **Communication knowledge test/34** |  |  |  |  |  |
| Site A (n = 30) | 28.0 | 29.0 | 1.0 | 0.1 to 1.9 | 0.02 |
| Site B (n = 17) | 26.4 | 26.4 | -0.1 | -1.5 to 1.4 | 0.9 |
| Site C (n = 17) | 29.3 | 29.8 | 0.5 | -0.5 to 1.5 | 0.3 |
| Total (n = 64) | 27.9 | 28.5 | 0.6 | 0.01 to 1.2 | 0.05 |
| **Confidence in Dementia Scale/45** |  |  |  |  |  |
| Site A (n = 30) | 31.7 | 38.3 | 6.6 | 4.5 to 8.7 | <0.001 |
| Site B (n = 19) | 35.6 | 39.7 | 4.1 | 2.1 to 6 | <0.001 |
| Site C (n = 17) | 32.9 | 40.4 | 7.5 | 5.7 to 9.3 | <0.001 |
| Total (n = 66) | 33.1 | 39.2 | 6.1 | 4.9 to 7.3 | <0.001 |

\* Based on paired t-tests

Staff reported a range of learning including: increased ability to identify agitation/distress; better understanding of individual patient behaviours and needs; how to respond using positive techniques they had learned, including entering the person’s reality, distraction, using the rules of conversation and providing explanations; improved ability to communicate with patients who are non-verbal; and practical, tailored de-escalation and diversion techniques for patients who are distressed.

> *“I learnt how to adapt my communication. Felt humbled and reflected on how I don’t always give patients the time that they deserve … Reflected on how taking an extra 5-10 minutes to explain assessment, purpose etc can actually facilitate the assessment in being more productive and effective”.* Staff survey response site C
>
> *“One particular group gave some really good examples of re-entering into someone’s reality, then using that to find distraction and they said it was a really positive experience and it worked.”* (Clinical Educator site A)
>
> *“Rules of conversation - I hadn’t considered this before how the lack of an answer could increase distress.”* (Staff survey site A)
>
> *“You don’t blatantly lie, you deflect and you’ve got to do that because if you know you’re not going to do it then it can have an adverse effect with the patient.”* (Staff participant site C)

Staff across all sites reported that the training validated their existing practices.

> *“I think it was nice to have a reinforcement of here are the things that we know work and, on the whole, will work more often than not. And the things that we know will only escalate the patient’s situation and get somebody more agitated”.* (Staff participant site C)
>
> *“I did find it useful, because it consolidates sometimes what you already know, even though you don’t realise that you know it sometimes.”* (Staff participant site A)
>
> Many staff also mentioned improved confidence in communicating with distressed patients.
>
> *“So I get [from the training] options of how to deal with a patient, if one strategy did not work, I will try the other one and then the other one again.”* (Staff participant site B)
>
> *“…one thing is the confidence of applying it. It just told me that what I’m doing is effective.”* (Staff participant site B)
>
> *“I have certainly seen staff that have attended the training being a lot more confident in how they are with dementia patients on the ward. Certainly, a couple of our TNAs [Trainee Nursing Associates] who’ve done the course, they said from the start, yes, it was brilliant, and you can see that confidence that they have now in what they’re doing.”* Ward manager site C

### Behaviour change

Ninety-one percent of participants said they planned to employ the learned skills regularly in practice. Staff across all sites highlighted the positive impact of understanding and finding a shared aspect of the patient’s reality, as well as the ways in which challenging someone’s reality or building a false reality might increase distress and be less effective

> *“One of the things in the training was like not to give false reality. You know when someone says where’s my son? And they might be like he’s coming this afternoon. Even though that person doesn’t know if they are, but it can be a quick statement when someone to just kind of use to calm, but it can affect, I think it can affect that trust that the patient has with someone because you are kind of feeding lies in a way.”* (Staff participant site C)
>
> *“Before … all the time I always bring them to the reality. Because I think in nursing school that’s what they taught us, to re-orientate the patients to time, place and reality. I didn’t know before that you could join them, and then explore a bit of what they’re [experiencing], because they could be in a different time of their life.”* (Staff participant site B)

Staff gave concrete examples of how they had applied training strategies into their practice including adapting the content of their communication to avoid escalation of distress, with practices such as acknowledging (not ignoring) patient’s repetitive or hard to understand comments and questions, offering structured redirection (for example to meaningful activities).

> *“There’s certain things that are definitely new … like the entering into the reality bits, …maybe it’s something that I would’ve tried but then I wouldn’t have known if that’s the right thing to enter into their reality.”* (Staff participant site A)
>
> *“… the lady that I went to, she was saying she wanted to go home. [Before] I’ll try to say to her, OK, if you have your dinner, you can go home, but I don’t do that anymore. I just ask her, have you spoken to anybody this morning in regards to going home? Stuff like that, so I haven’t found myself going back to it, because from the training I see that it’s not really fair on them and it doesn’t work.”* (Staff participant site C)
>
> *“I had a patient who was saying she is giving birth, but she’s already 76. Then when I explored that reality, I knew that … she was actually having abdominal pain, so she was thinking that the abdominal pain is her giving birth … It was actually really helpful to find out why she was acting like that.”* (Staff participant site B)
>
> *“The way they [staff] are using the words, and also the posture and the positions*
>
> *… how they’re facing the patient. Things like that, you could see there is difference, so the whole approach.”* (Ward manager site A)
>
> *“that was quite a big learning point for me that people with dementia will know when you’re just essentially ignoring them. And if you don’t respond then they will feel more annoyed which makes so much sense. But so I’m very careful now. If someone is saying something, and even if it’s repetitive or it’s not really understandable to me, if I’m at least kind of acknowledging it. … and I find myself thinking, oh, I’m, I’m putting my training into good use today.”* (Staff Participant Site C)

Although some staff raised challenges of applying the general techniques to the specific needs of patients they cared for.

> *“After the class we tried to incorporate some of the ideas we got from the class with our patients. Still it’s difficult because each patient is different, we can’t apply the same thing with the different patients, it will work differently.’* (Staff participant site B)

The observations of communication behaviours in practice showed increases in the mean number of positive practices observed per 100 minutes post-training in two sites, and a reduction in the third site (see Table 5). There was a low occurrence of negative practices at baseline, and this increased slightly in two sites and reduced in the third. However, these data should be treated with caution as there was variation in the overall number of observation minutes between sites and at each time point, the same numbers of staff and the same staff members were not observed at each time point.

**Table 5:** Observational outcomes data.

| Scale<br>(n= pre, post) | Pre-<br>training<br>(T0) | Post-training<br>(T2) | Mean<br>difference | 95% CI | p-<br>value* |
| --- | --- | --- | --- | --- | --- |
| ***Positive communication practices (mean per 100 mins observation)*** |  |  |  |  |  |
| Site A | 2.4 | 1.3 | -1.1 |  |  |
| Site B | 0.4 | 1.1 | +0.7 |  |  |
| Site C | 2.5 | 5.8 | +3.3 |  |  |
| Total | 1.8 | 2.7 | +0.9 |  |  |
| ***Negative communication practices (mean per 100 mins observation)*** |  |  |  |  |  |
| Site A | 1.3 | 0.4 | -0.9 |  |  |
| Site B | 0 | 0.3 | +0.3 |  |  |
| Site C | 0.4 | 0.5 | +0.1 |  |  |
| Total | 0.6 | 0.4 | -0.2 |  |  |
| ***CMAI-O/29*** |  |  |  |  |  |
| Site A<br>(n=13,14) | 30.4 | 32.6 | 2.3 | -1.4 to 5.9 | 0.2 |
| Site B<br>(n=15,12) | 33.5 | 30.8 | -2.8 | -7.1 to 1.5 | 0.2 |
| Site C (n=8,6) | 37.1 | 30.7 | -6.5 | -13.8 to -0.9 | 0.08 |
| Total (n=36,32) | 33.2 | 31.6 | -1.6 | -4.4 to 1.1 | 0.2 |
| ***PAS mean/4*** |  |  |  |  |  |
| Site A<br>(n=13,14) | 2.1 | 2.0 | -0.1 | -1.1 to 1.2 | 0.9 |
| **Site B**<br>**(n=15,12)** | 1.8 | 2.5 | 0.7 | -0.9 to 2.3 | 0.4 |
| **Site C (n=8,6)** | 3.5 | 1.8 | -1.7 | -5.1 to 1.7 | 0.3 |
| **Total (n=36,32)** | 2.3 | 2.2 | -0.1 | -0.9 to 1.10 | 0.8 |

\* Based on non-paired t-tests

### Outcomes for patients, families and services

The observational data of patient agitation (Table 5) indicated a slight reduction in agitation intensity and frequency overall, also reflected in individual data at two of the sites. However, this was not statistically significant, and the data must be treated with caution given the small sample size and that different patients were observed at T0 to T1.

Data on systemic outcomes from the training, for example through reports of reduced incidents or cost savings were not evidenced in the interviews. A range of smaller scale positive impacts was identified. For example, staff across all sites noted patients seemed calmer and there appeared to be fewer escalations of distress.

> *“When the patient was confused and trying to get out from the ward. He is trying the fork and spoon to open the door, then I asked him what was his job? Then he was more settled and he was telling me about his job, and when he was mechanical engineer … then [for] nearly one, two-and-a-half hours he sat near me, and he was talking, then that made him calm down.”* (Staff participant site A)
>
> *“The ward environment is calm, everybody is, because they’re very disturbed sometimes.”* (Staff participant site B)

Staff in all sites felt the training had supported some change in staff culture, with staff at site B applying dementia-friendly principles to all patients, and in sites A and C a greater emphasis in team approaches on reflection on patient experiences and realities was noted. Some staff members commented on using their new skills to support colleagues who had not accessed the training, in communicating with patients.

> *“I speak with colleagues, and also with those who can’t attend…we’ve still got some new staff in the ward, so we told them that we can use this kinds of techniques while dealing with a dementia patient, it was really helpful for them also.”* (Staff participant site A)
>
> *“So, that’s where I stepped in earlier and tell that colleague, “You want to try doing this instead,” and that got her [the patient] quite settled for a long time.”* (Staff participant site C)
>
> *“I told my husband as well, he is working in admissions department, so he didn’t have the training. I told him if the patients come in like this, just try this one.”* (Staff participant site A)

In all sites relatives commented on staff attentiveness and use of a personalised communication approach including some of the trainable techniques, in this case offering explanations.

> *“The treatment and communication are better in here … they have a lot of people with dementia and talk to them in a way that makes sense”* (Family member site A)
>
> *“We could hear they were talking to him, just letting him know what they were going to be doing, which was good.”* (Family member site C)

In summary, the training was challenging to organise, largely the most motivated staff attended, and fewer staff were trained than anticipated in two sites. Training was well received by participants for its content and delivery methods. It led to some observed and reported improvements in staff’s communication knowledge, and confidence to communicate with people living with dementia. Staff reported a range of ways in which the training had impacted changes in their communication approaches and indicated that they felt the training had led to reductions in patient distress and improvements in staff practices. However, these were challenging to evidence objectively through quantitative measures of communication practice and patient outcomes. Some barriers to implementation were identified ranging across training delivery to putting communication behaviours into practice consistently.

## Discussion

This study evaluated the implementation of a communication skills training programme to help acute hospital staff to better respond to patients with dementia who may become distressed. As one of the few studies to date that has focussed on specifically addressing distress and related communication, it adds to current knowledge of how acute hospital staff can be best supported to deliver better care in this complex area.

Existing guidance on best practice in the design and delivery of dementia training underpinned the programme’s development likely resulting in the positive staff reactions to the training. Specifically, face-to-face, interactive training that includes a variety of practical learning resources that support application of theory into practice, and opportunities for discussion and reflection (30–32, 43). Of note in our training was the inclusion of real-world video recordings collected in the phase one Conversation Analysis study, which participants identified as particularly impactful for learning. The efficacy of video content that depicts real-world scenarios is well documented in pre-registration nurse education for supporting learning, problem solving and skills development (44, 45). This literature, however, identifies the resource issues and challenges in creating video content that is authentic(45). The value of video data collected for Conversation Analysis in providing authentic content to underpin training in healthcare contexts has been highlighted(24, 46). This study adds to the growing body of evidence on the impact of utilising evidence-based principles in training design and delivery and of authentic video-based resources to support staff learning and its implementation into practice.

Despite staff scoring highly on the communication knowledge measure at baseline and only a slight increase in knowledge post-training, in the interviews staff reported high training relevance, good knowledge gains across a range of areas, and increased communication skills confidence. These findings mirror those of our previous research implementing a Conversation Analysis-based general communication skills training programme in acute hospitals(38). One explanation for this is that our self-developed communication knowledge measure was inadequate for measuring knowledge gained from the training. However, it may also be the case that because Conversation Analysis is based on real-world interactional practices, staff already have knowledge on best practice approaches to good communication with people with dementia, but may be unaware of, or lack confidence in, which approaches to try and which to avoid. The role of Conversation Analysis in illuminating existing positive practices and providing a rationale for their application has already been documented by our team(47), and this study adds to our understanding of this phenomenon. Conversation Analysis may be particularly helpful in complex communication scenarios like distress when approaches may not produce tangible outcomes in patient behaviour. For example, if a communication approach prevents escalation rather than stops distress it may be difficult for a staff member to understand if, or how, their approach has been beneficial. The use of Conversation Analysis can, therefore, provide an evidence-based framework and practical toolkit of positive (and negative) communication approaches to underpin staff’s application of existing skills and knowledge to complex practice situations.

Whilst the two half-days of training was challenging for the clinical educators to arrange, this format was received positively by staff who welcomed the opportunity to assimilate the information, reflect and apply approaches in practice between sessions. Kolb’s experiential learning cycle(48) provides a theory for translation of concrete experiences through a process of reflection, to abstract conceptualisation and then active experimentation in practice, which has been widely applied in healthcare education(49, 50). Reflection has also been identified as playing an important role in health professional practice confidence(51). Thus, our adoption of reflection and application time within the programme structure may have contributed to the increase in staff confidence we evidenced. Therefore, future delivery of the programme will need to consider balancing the practical aspects of scheduling two half-day sessions against the potential value this format brings for learning and its application into practice.

However, despite the programme being underpinned by this cycle, some staff indicated moving from the video and transcript-based concrete examples of communication approaches, to abstract application in a range of alternative practice scenarios, remained challenging. Therefore, it may be further support is required, for example through mentorship or coaching, and further opportunities for discussion with peers(52), for some staff to engage successfully in the stages beyond concrete experience and to apply learning successfully into practice. Informally the clinical educators have reported plans to continue delivery of the training in multiple shorter sessions, which may allow increased opportunities for discussion, reflection and implementation between each session.

Whilst staff reported a range of impacts of the training on their own practice, and more limited wider impacts on overall practice, obtaining objective assessment of the impact of training on staff communication behaviours and particularly on patient outcomes was challenging. Previous reviews have identified the heterogeneous nature of data collection methods and outcome measures utilised in evaluations of communication training and have subsequently questioned the robustness and thus clinical relevance of findings(53). The nature of acute hospital settings with a wide range of staff providing patient care and high throughput of patients alongside the necessity to adapt communication approaches to situations and individual patient needs, presents specific evaluation challenges. This may account for why most evaluations of dementia training in acute hospitals have failed to include robust evaluation of impact on staff practice behaviours or patient outcomes(30, 54). Our study has further highlighted the limitations of using quantitative measures alone to evaluate practice and patient focussed outcomes of training. For example, observational measures of agitation/distress may be utilised to overcome potential biases caused by different staff completing proxy measures(39), however acute hospital environments introduce a range of additional complexities to quantitative comparison of pre-post observational agitation data. Our study researchers noted distinct environmental conditions that could have influenced agitation levels at each site and across time points, independent of the training intervention. For example, differences in patients (e.g. how acutely unwell and/or how much time was spent asleep, one particularly distressed patient impacting the well-being of others), the numbers of visitors (with visitors providing stimulation and reducing the likelihood of patient distress), the length of mealtime periods included in observations (where people were engaged and less likely to be distressed), environmental differences (a ward move pre to post training, malfunctioning air-conditioning unit impacting ward temperature, positive spaces such as day rooms) and staffing levels in pre-versus post-training observation periods. Our use of case study methodology permitted multiple sources to be drawn on to gain a more nuanced understanding of training impact across all four Kirkpatrick levels. While limitations to understanding impacts of our training on staff practice and patient outcomes remain, we have been able to provide some evidence across all levels. Thus, case study methodology may be a more appropriate methodology to evaluate complex training interventions such as communication skills in acute hospital settings, than randomised controlled trials or quasi-experimental designs.

### Limitations

The quantitative findings must be treated with caution since sample sizes lack statistical power and observation periods were not comparable (time, staff or patients) across time points. Staff measures only included data from respondents who completed the measure at both time points. Contextual factors must be considered when interpreting agitation scores across sites. Whilst some staff had received training, post-training observations were conducted at times when untrained staff were on duty, and it is therefore unlikely that the decrease in scores can be attributed to the intervention alone.

## Conclusions

It is feasible, although challenging, to deliver empirically based communication skills training to support acute hospital staff to better care for people living with dementia who may become distressed. It can lead to staff perceived increases in knowledge and confidence to support distress in this population. Utilisation of best practice guidance in training design and delivery facilitated high levels of staff satisfaction with the programme, which is important for sustainable implementation. Future research should consider ways in which impacts on patient outcomes can be evaluated.

## Data Availability

All data produced in the present study are available upon reasonable request to the authors

## Acknowledgements

We would like to thank the participating NHS Trusts, their staff, patients and families for taking part in this study. We would also like to thank, Lauren Bridgstock, Sarah Field-Richards, Katarzyna Kowalewska, Fran Labrom, Amy Love, Sherena Nair, Sean Ninan, Alison Raycraft, Lovelyn Umeloh, Cas Shotter Weetman, Isabel Windeatt-Harrison for their support with the study.

## Author contributions

O’Brien, Rebecca – Conceptualization, Data curation, Formal analysis, Funding acquisition, Investigation, Methodology, Project administration, Supervision, Writing – original draft

Theodosopoulou, Danai - Data curation, Formal analysis, Investigation, Project administration, Writing – original draft

Janes, Marie - Data curation, Formal analysis, Investigation, Writing – original draft Clark, Rachel - Data curation, Formal analysis, Investigation, Writing – original draft

Harwood, Rowan – Conceptualization, Formal analysis, Funding acquisition, Methodology, Project administration, Supervision, Writing – original draft

Papworth, Andrew – Investigation, Writing – review and editing

Beeke, Suzanne – Conceptualization, Funding acquisition, Methodology, Project administration, Supervision, Writing – review and editing

Bramley, Louise – Funding acquisition, Project administration, Writing – review and editing

Chivinge, Aquiline – Funding acquisition, Project administration, Writing – review and editing

Goldberg, Sarah – Conceptualization, Funding acquisition, Methodology, Project administration, Supervision, Writing – review and editing

Haines, Sue – Funding acquisition, Project administration, Writing – review and editing

Pilnick, Alison – Conceptualization, Funding acquisition, Methodology, Project administration, Supervision, Writing – review and editing

Sartain, Kate - Funding acquisition, Project administration, Writing – review and editing

Whitworth, Morag - Project administration, Writing – review and editing

Surr, Claire – Conceptualization, Formal analysis, Funding acquisition, Methodology, Project administration, Supervision, Writing – original draft

